# A decade of dengue disease burden in Africa (2013–2023): a systematic review

**DOI:** 10.1101/2024.10.22.24315827

**Authors:** Gaspary O. Mwanyika, Monika Moir, Abdualmoniem O. Musa, Jenicca Poongavanan, Graeme Dor, Eduan Wilkinson, Cheryl Baxter, Tulio de Oliveira, Houriiyah Tegally

## Abstract

**Background:** Dengue is a major mosquito-borne disease worldwide. The epidemiological trends of the disease in Africa over the past decade remain unclear. This review aims to provide insight into the epidemiological trends of dengue in Africa from 2013–2023.

**Methods:** We systematically searched PubMed/MEDLINE and Scopus for studies published between January 2013 and December 2023. Additionally, we collected official records from the World Health Organization for Africa and African Centre for Disease Control. We included studies that reported dengue cases in humans in Africa and excluded publications prior to 2013, review articles and non-human studies. For specific countries, the suspected cases per 100,000 population and fatality rates were estimated and the trend predicted using a negative binomial model. The statistical analyses and visualisations were performed using R programming.

**Results:** Of the 453 reports screened, 87 from 25 African countries were selected for systematic review. Between 2013 and 2023, approximately 200,000 suspected dengue cases, 90,000 confirmed cases and 900 deaths were reported in Africa. Over 80% of confirmed cases originated from West Africa, with Burkina Faso reporting over 500 cases per 100,000 population. DENV1 and DENV2 predominating at different times with transmission closely linked to rainy seasons.

**Conclusions:** The rising dengue cases across Africa, highlight the need to strengthen surveillance and implement effective regional-specific interventions against future dengue outbreaks. Further research is necessary to improve our understanding on dengue transmission dynamics and suitability of regions in Africa.

## 1. Introduction

Dengue is a major mosquito-borne viral disease transmitted by infected female mosquitoes of the *Aedes* genus [1]. The disease is caused by dengue virus (DENV) of the Flaviviridae family in the genus *Orthoflavivirus.* The virus consists of four antigenically distinct serotypes: DENV1, DENV2, DENV3 and DENV4, which are capable of inducing mild to severe illnesses in humans [2]. Globally, approximately 390 million dengue infections, 500,000 hospitalizations and more than 20,000 deaths are reported annually [3]. In recent decades, the virus has transcended its traditional boundaries, extending into temperate regions including Europe and North America [4,5]. The disease has been reported in Africa since the late 19^th^ and early 20^th^ centuries, with reported cases in Zanzibar (1870), Burkina Faso (1925), Egypt (1887), South Africa (1926–1927), and Senegal (1927–1928). From the 1960s to 2010, laboratory confirmed cases were reported in 15 African countries, with recent endemicity established in more than 34 countries [6]. Although the prevalence of dengue virus across Africa has been previously documented [7,8], epidemiological trends in the context of morbidity and mortality, geographical distribution, seasonality and transmission suitability remain unclear. In this systematic review, we collate a decade of dengue epidemiological data from the African continent to synthesise and analyse epidemiological trends between 2013 and 2023. We highlight regional and country-specific epidemiological trends to inform public health response against future dengue outbreaks in Africa.

## 2. Methods

### 2.1 Search strategy and selection criteria

This review analysed studies and reports describing dengue cases in African countries and territories. Medical Subject Headings (MeSH) terms such as: dengue virus, prevalence, epidemiology, serotype, in conjunction with a compilation of specific African countries and territories were used in the search. We searched PubMed/MEDLINE and Scopus databases for relevant English articles using an advanced search strategy (Appendix S1). Additional epidemiological reports were obtained from the World Health Organization for Africa (WHO AFRO) and the African Centre for Disease Control (AFRICA CDC) between January 2013 and December 2023. We included articles that reported dengue cases in human studies in Africa and official records from the WHO AFRO and AFRICA CDC surveillance reports. We excluded publications prior to 2013, review articles, and non-human studies.

### 2.2 Screening and quality assessment

Two reviewers (GOM and AM) conducted an initial screening of the titles and abstracts from the search results using Rayyan application software accessible at https://rayyan.ai/ to identify relevant articles. Reasons for exclusion of irrelevant articles were documented. Three reviewers (GOM, AM and HT) evaluated the quality of included reports. The selection of reports for inclusion in the systematic review followed the Preferred Reporting Items for Systematic Reviews and Meta-analysis guidelines (PRISMA) [9].

### 2.3 Key definitions

The following terms have been used in the analysis to signify different metrics used to describe dengue burden in Africa.

1. A suspected case was defined as any individual residing in or having travelled to areas with dengue transmission within the past 14 days and presents with acute fever, typically lasting from two to seven days’ duration with two or more of the following symptoms: nausea/vomiting, abdominal pain, chills, rash, headache/retro-orbital pain, myalgia and arthralgia; may exhibit petechial or positive tourniquet test (+ >10 pinpoint-sized spots of bleeding under the skin (petechiae) per square inch, low platelet and white blood cell counts even without any warning sign.
2. A confirmed case was defined as a suspected dengue case with laboratory confirmation of infection which may include polymerase chain reaction (PCR), virus culture, IgM seroconversion in paired sera (acute and convalescent samples), IgG seroconversion in paired sera or fourfold IgG titre in paired sera.
3. A severe case was defined as a suspected/probable/confirmed dengue case presenting with one or more of the following symptoms: severe plasma leakage leading to dengue shock syndrome fluid accumulation with respiratory distress, severe bleeding, severe damage of organs such as liver (aspartate aminotransferase (ASAT) or alanine aminotransferase (ALT) elevation ≥ 1000) and central nervous system.
4. Case fatality rate was defined as the proportion of deaths within a specified population that are attributable to the total number of suspected dengue cases over a specific period.
5. Transmission potential (index P) was defined as a measure that quantifies the risk of dengue virus transmission in a specific region, taking into account climate-based factors such as temperature and humidity, which directly influence the breeding, survival, and biting behaviour of the mosquito vectors responsible for spreading the virus. This index provides insight into how favourable environmental conditions are for dengue circulation in a given area. The index was developed by Nakase et. al (2023) [10]. The spatio-temporal estimates of transmission potential were then compiled for African countries and territories. A threshold of 1.0 was selected to compare with the basic reproductive rate. A value of 1.0 indicates that in a population where the average number of adult female mosquitoes per host is 1.0 corresponds to a reproduction number of 1. The period of transmission suitability is defined as a month in which the transmission potential is greater than 1.0 [10].

### 2.4 Data synthesis and analysis

Data on the year of outbreak, country of origin, number of suspected and confirmed cases, deaths and serotype counts were extracted from included reports and compiled into an Excel spreadsheet (Miscrosoft Corp., 2016 Redmond, WA, USA). The estimation of dengue burden was based on the number of suspected cases per country’s population during the respective years. The population data for the respective years were obtained from the Worldometer web-source, accessible at https://www.worldometers.info/population/africa/. Case fatality rate (CFR) was computed based on the number of reported deaths per total number of suspected dengue cases in the specific country. A negative binomial model was used to predict the growth of suspected cases using the year as a predictor variable. The model was selected to suit count data, such as dengue case numbers and accounts for overdispersion. The model was presented by the following equation;

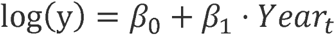

Where;

y = Expected number of suspected dengue cases

*β*_0_ = Intercept of the model

*β*_1_ = Coefficient for the year t

*Year_t_* = The year variable for which the number of dengue cases was predicted

*e*^*β*0+*β*1^ = A multiplicative factor for the growth in dengue cases. The best fit was compared with a Poisson regression model using likelihood ratio test. Statistical analysis and visualizations were conducted using R version 4.3.2 with primary packages ggplot2, dplyr and MASS.

## 3. Results

### 3.1 Literature search

The review protocol was registered in the PROSPERO International prospective register for systematic reviews of human studies under CRD42023480486. The search yielded a total of 453 results, including 297 from academic databases and 156 official sources. After removing 151 duplicates, 302 unique records were screened by titles and abstracts. 173 reports were excluded from the screening process, and 129 were evaluation for eligibility. Finally, 42 records were excluded for specific reasons (Appendix S2), and 87 included in the systematic review (**Figure 1**).

**Figure 1.**
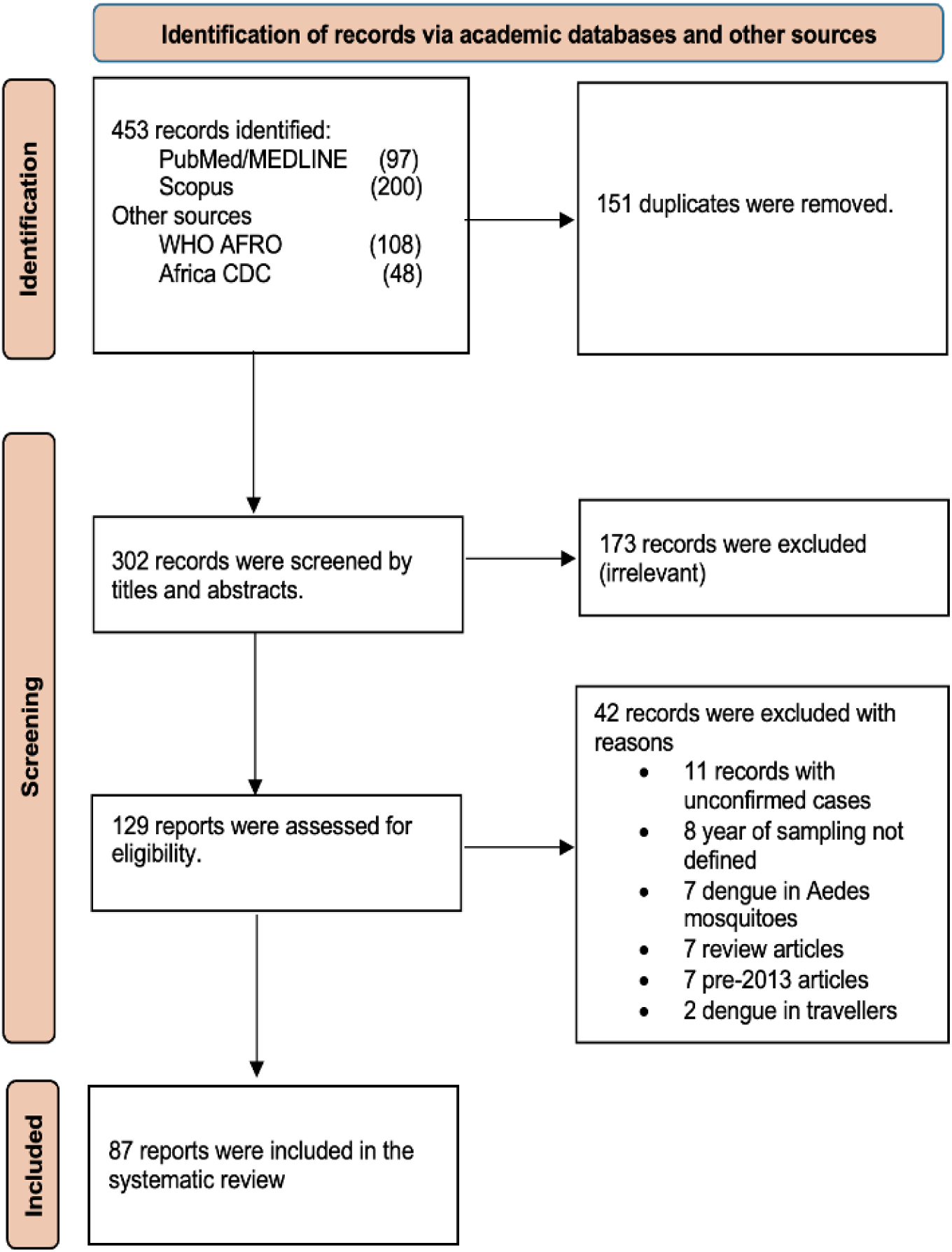
Selection process of included reports according to PRISMA guideline.

### 3.2 An overview of included reports

Table 1 presents the epidemiological data extracted from the included reports, comprising 35 published articles and 52 official records from the WHO AFRO and AFRICA CDC surveillance data. Between January 2013 and December 2023, data on dengue were available from 25 African countries. Seven countries, including, Burkina Faso (7 records, n = 14,7286), Cameroon (6, n = 2513), Cote d’Ivoire (5, n = 8,250), Kenya (8, n = 7,828), Senegal (8, n = 4,606), Sudan (7, n = 2,696) and Tanzania (6, n = 10,369) accounted for more than 50% of all records (46/87) and 80% of cumulative suspected cases (19,7707/24,4352). A comparison year-on-year reveals that the highest number of reports (21.8%, 19/87) and suspected cases (60.3%, 174,888/245,143) were documented in 2023. In this year, Burkina Faso reported the most lethal outbreak with a total of 147,286/174,888 (84.2%) cumulative suspected cases, 68,402/72677 (94.1%) confirmed cases, and 688/765 (89.9%).

**Table 1.**
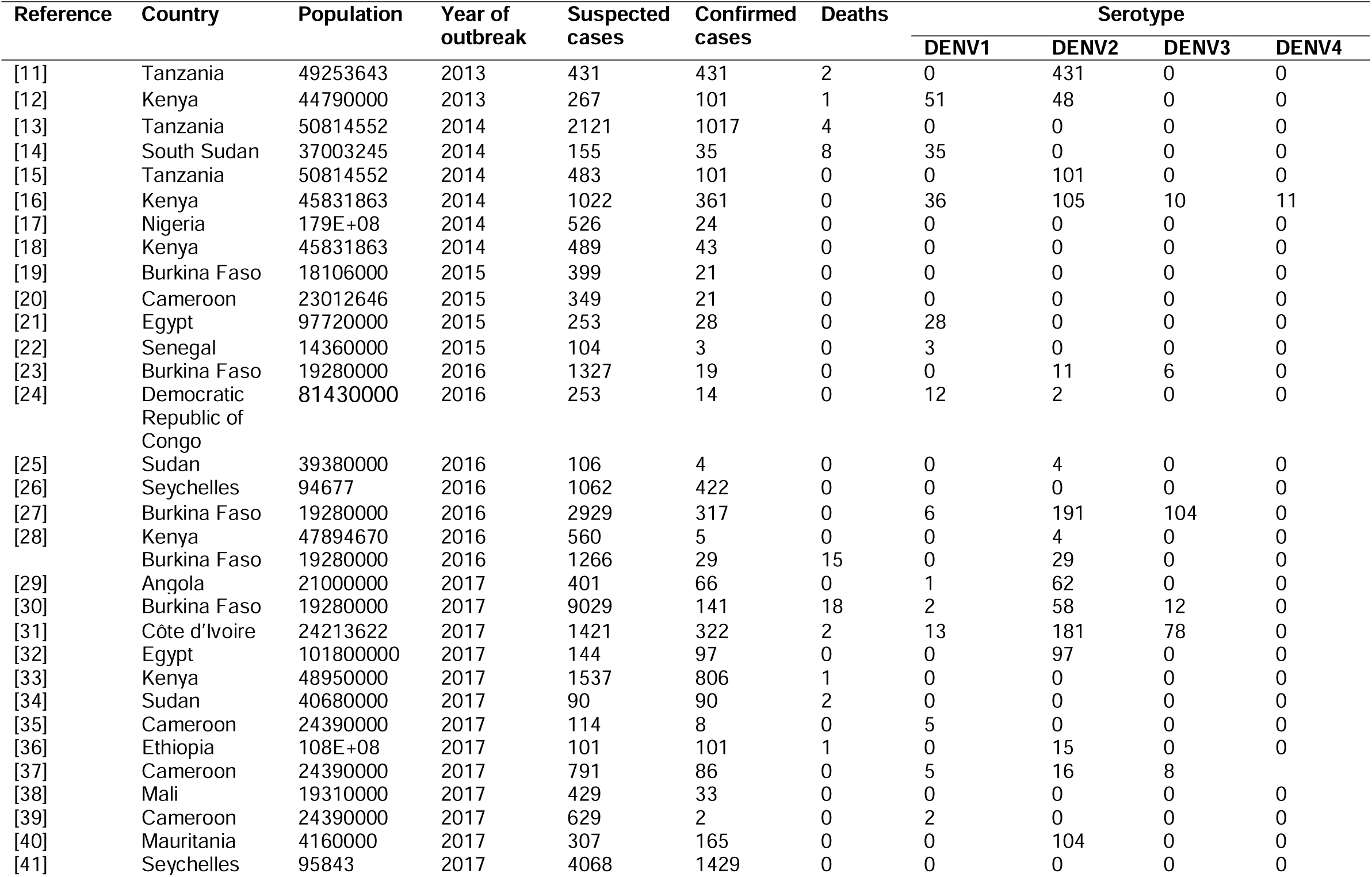
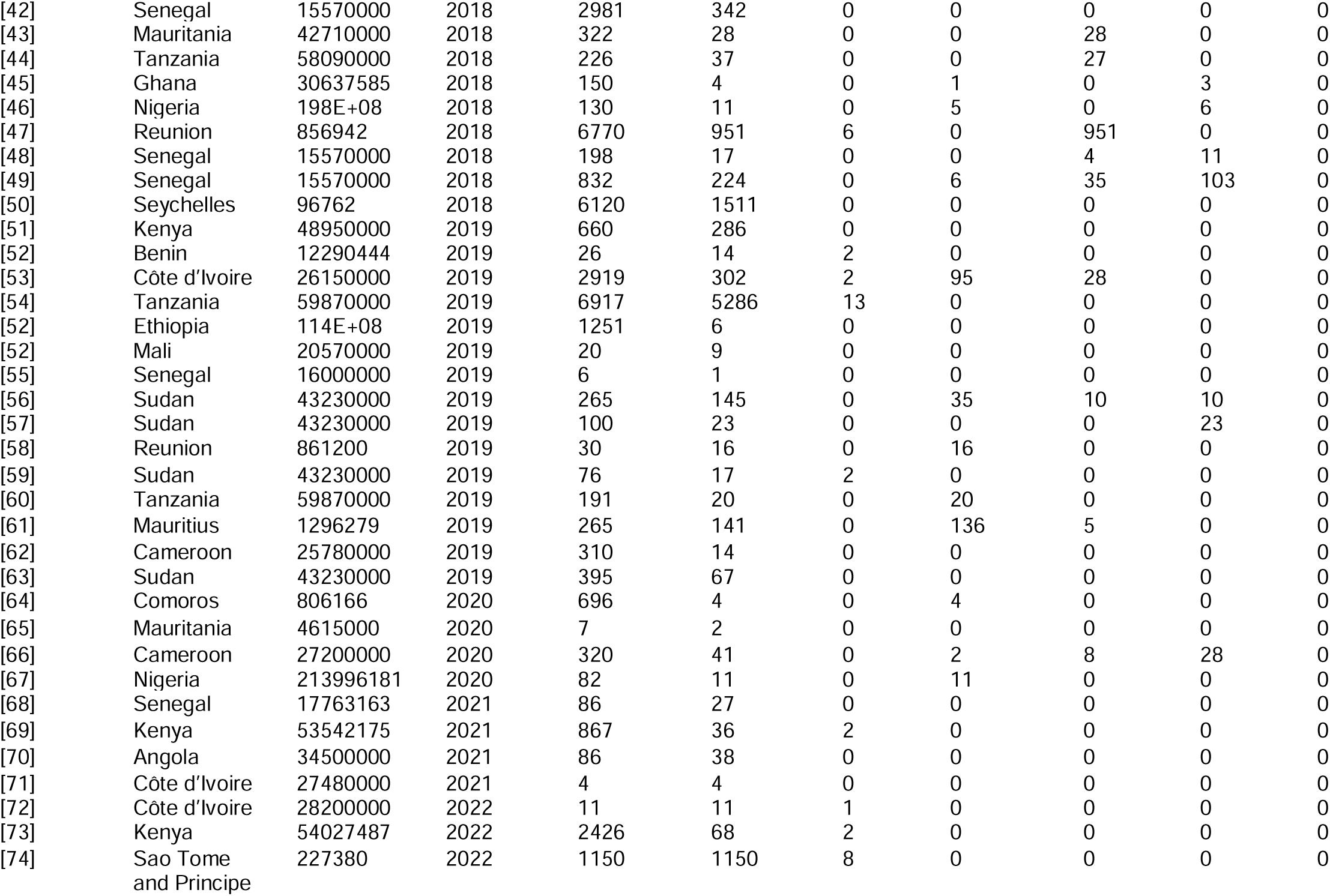
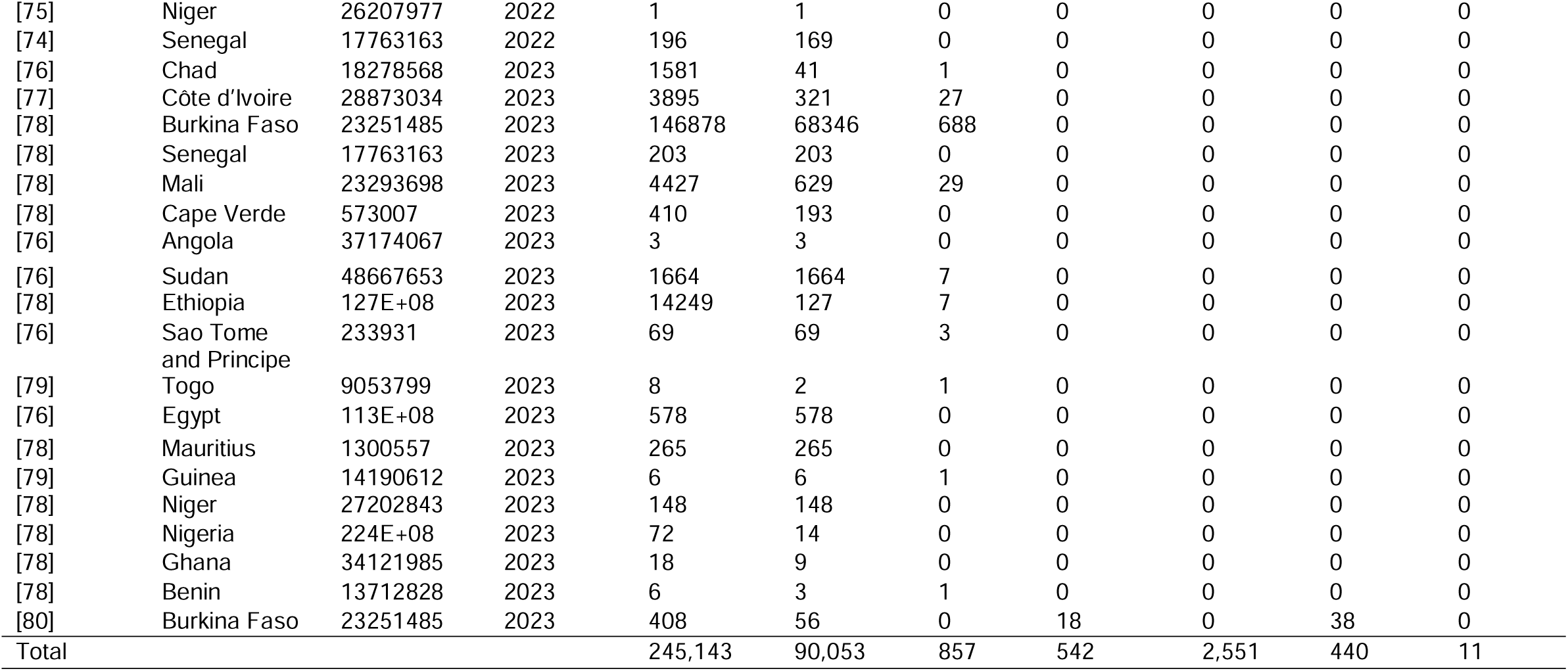
Epidemiological trends of dengue cases and deaths in Africa over the last decade (2013−2023, n= 87)

### 3.3 The spatial distribution of suspected dengue cases in Africa

Spatial distribution analysis indicates differences in number and distribution of suspected dengue cases across African countries and territories. From 2013-2023, Burkina Faso, Ethiopia and Tanzania reported the highest number of cases, surpassing 10,000. Reunion and Seychelles reported the highest number of cases (5,001––10,000) among territories (**Figure 2**).

**Figure 2.**
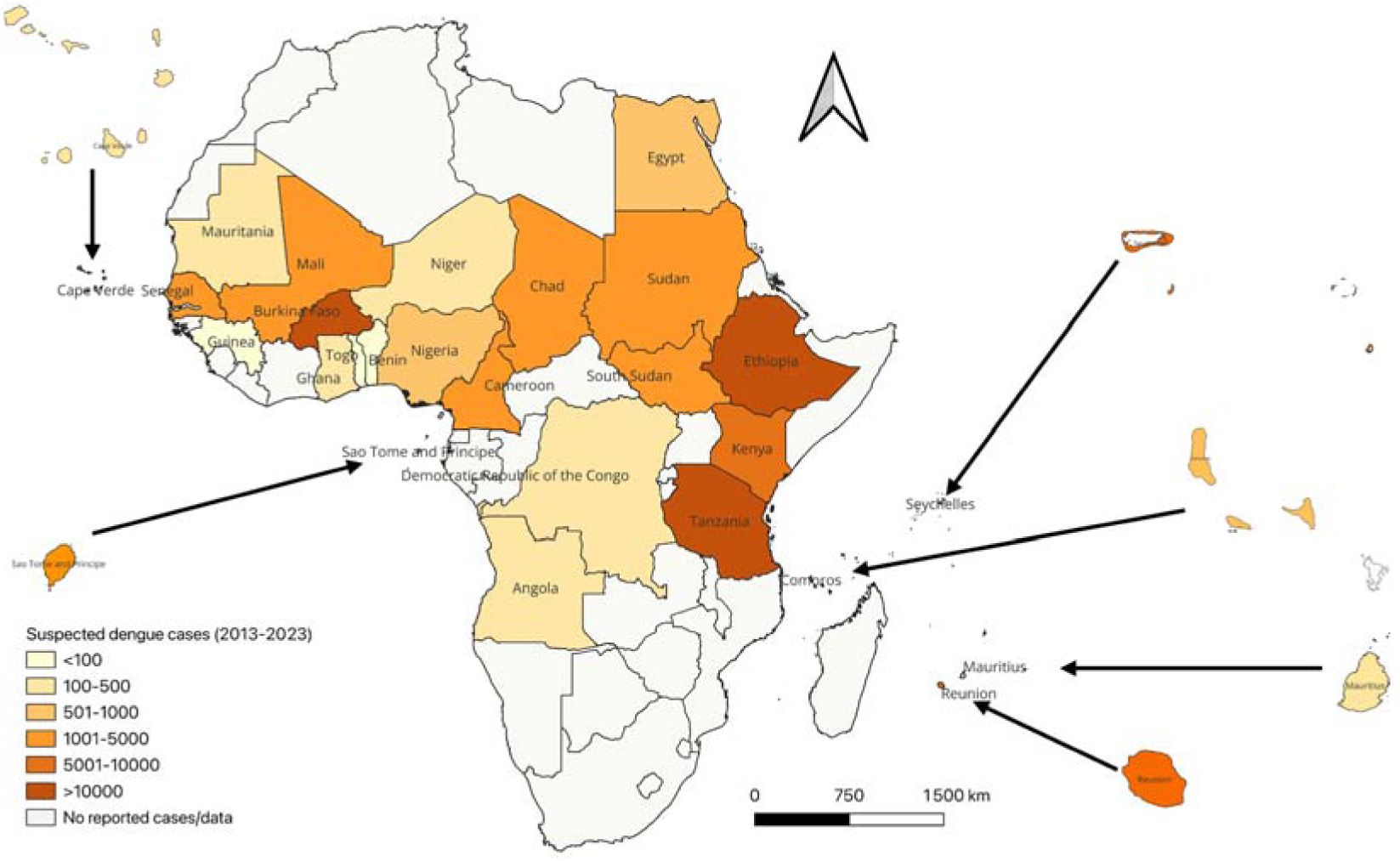
A map of Africa illustrating the geographical distribution of suspected dengue cases in various African countries and territories based on data reported from 2013-2023. The map was developed using QGIS open-source software version 3.38 accessed at https://qgis.org/download/

### 3.4 A rise in the number of confirmed dengue cases in West Africa

Over the past decade, dengue disease burden has increased in Africa, with a 5-fold increase in West Africa, from approximately 14,000 in 2014 to 70,000 cumulative confirmed cases in 2023. The region was responsible for approximately 80% of the confirmed cases (71, 793/89,967) (**Figure 3**).

**Figure 3:**
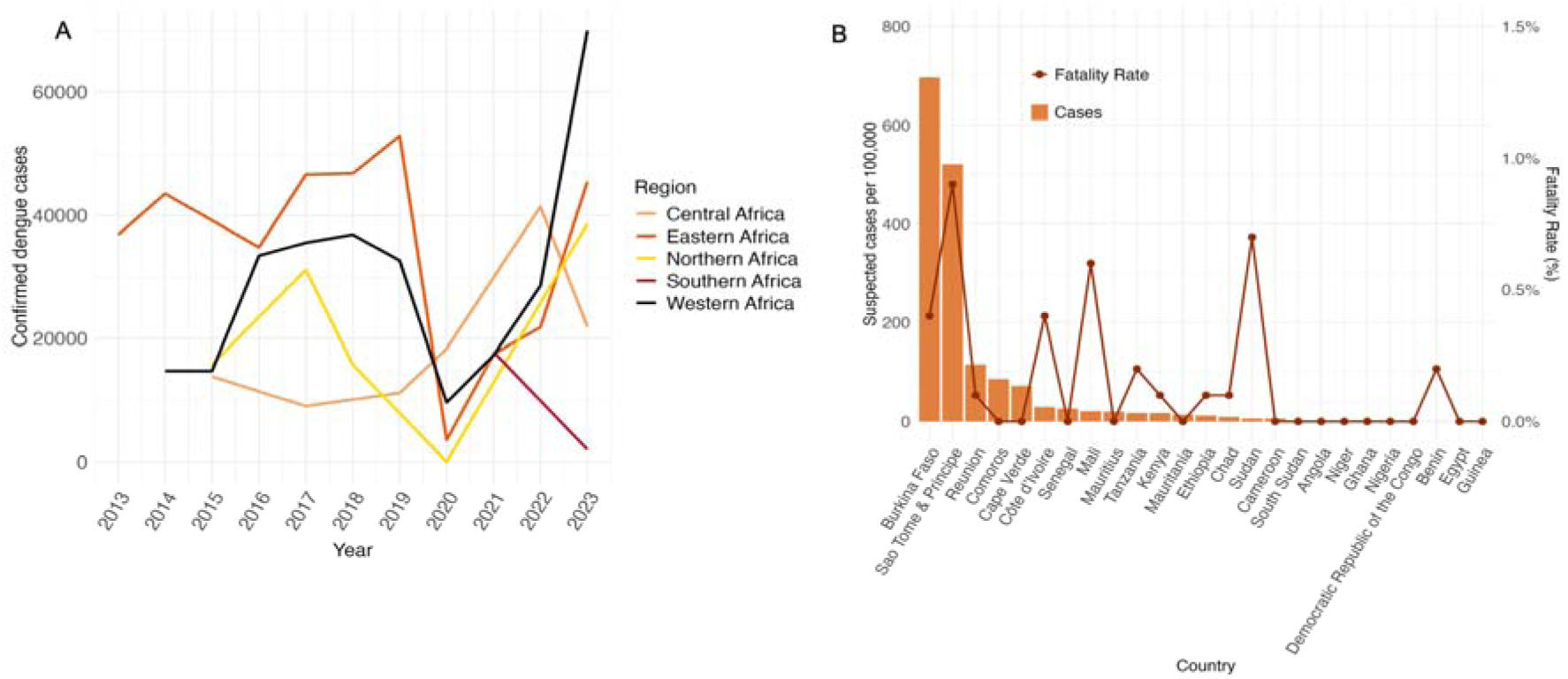
A. The magnitude and trend of cumulative confirmed dengue cases across Africa. B. The number of suspected dengue cases per 100,000 population and fatality rates (%) for specific countries based on data reported from 2013 to 2023.

Burkina Faso, and Sao Tome and Principe recorded over 500 cases per 100,000 population surpassing all other countries (Fig 3B). Sao Tome and Principe, Sudan and Mali reported higher fatality rates of 0.9%, 0.7% and 0.6% respectively.

### 3.5 The occurrence of multiple DENV serotypes and severe dengue

Since 2013, all four serotypes of DENV, DENV1, DENV2, DENV3, and DENV4) have been reported in Africa. Continentally, DENV1 and DENV2 dominated at different time (**Figure 4**). From 2019 to 2020, DENV1 was predominate serotype in both Eastern and Western Africa. In 2023, DENV3 dominated Western Africa whereas DENV2 prevailed in Eastern Africa.

**Figure 4.**
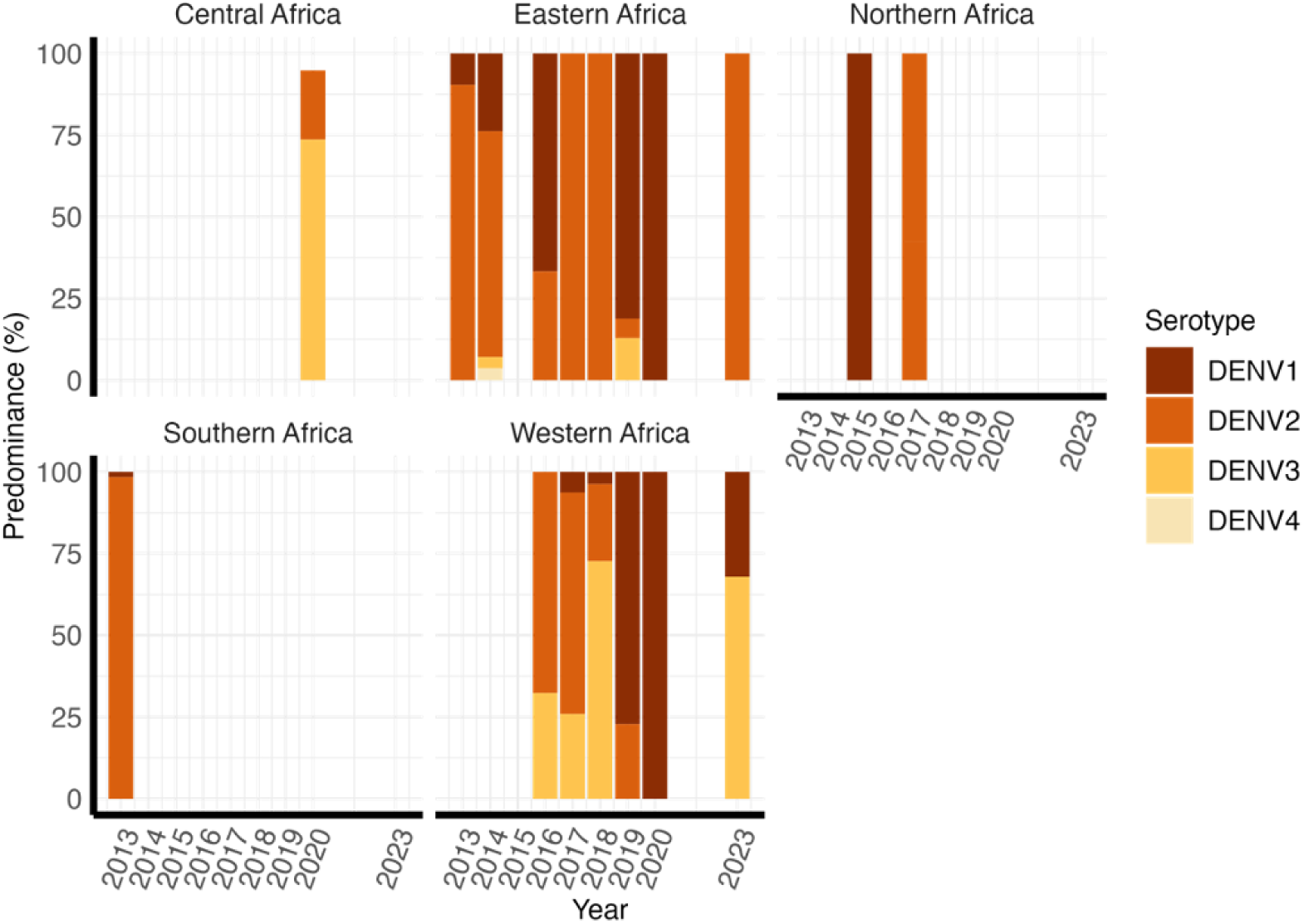
Spatiotemporal distribution of DENV serotypes in Africa based on the data available from 2013 to 2023.

Since 2013, a few cases of severe dengue, specifically Dengue Haemorrhagic Fever (DHF) and Dengue Shock Syndrome (DSS), have been reported in Africa. Twenty cases were reported in the United Republic of Tanzania (2014), nine in Burkina Faso (2015), five in Ethiopia (2017), two in Benin (2019) and 40 in Sudan (2019). Severe outcomes were associated with diabetes in Tanzania [11], pregnant women in Burkina Faso [19], male gender in Ethiopia [36] and malaria-dengue co-infection in Sudan [81].

### 3.6 Regional differences in transmission seasonality and suitability

Central (**Figure 5A**) and Eastern (**Figure 5B**) Africa experienced prolonged dengue transmission seasons from April to November. In Central Africa, high peaks were observed in June (> 400 cases), September (> 1,000) and November (> 700) with a monthly median of 94 cases. Eastern Africa exhibited a dynamic transmission pattern with the highest peaks in May and June (> 4,000), September and November (> 8,000, respectively) with a monthly median of 88 cases. Northern Africa (Figure 5C exhibited sporadic transmission patterns with high peaks in February, July, August and November (>200, respectively) with a monthly median of 93 cases. Western Africa (**Figure 5D**) had distinct high transmission seasons with high peaks in October (> 17,000), November (> 50,000) and December (30,000) and a monthly median of 98 cases while Southern Africa reported less than 100 cases. Western Africa reported the highest number of deaths (> 500), followed by Eastern Africa (> 40). In Western Africa, deaths were mostly reported during the high transmission season between October and December, whereas in Eastern and Central Africa, deaths were reported during both the low and high transmission seasons. There have been no reported fatalities in Northern Africa.

**Figure 5.**
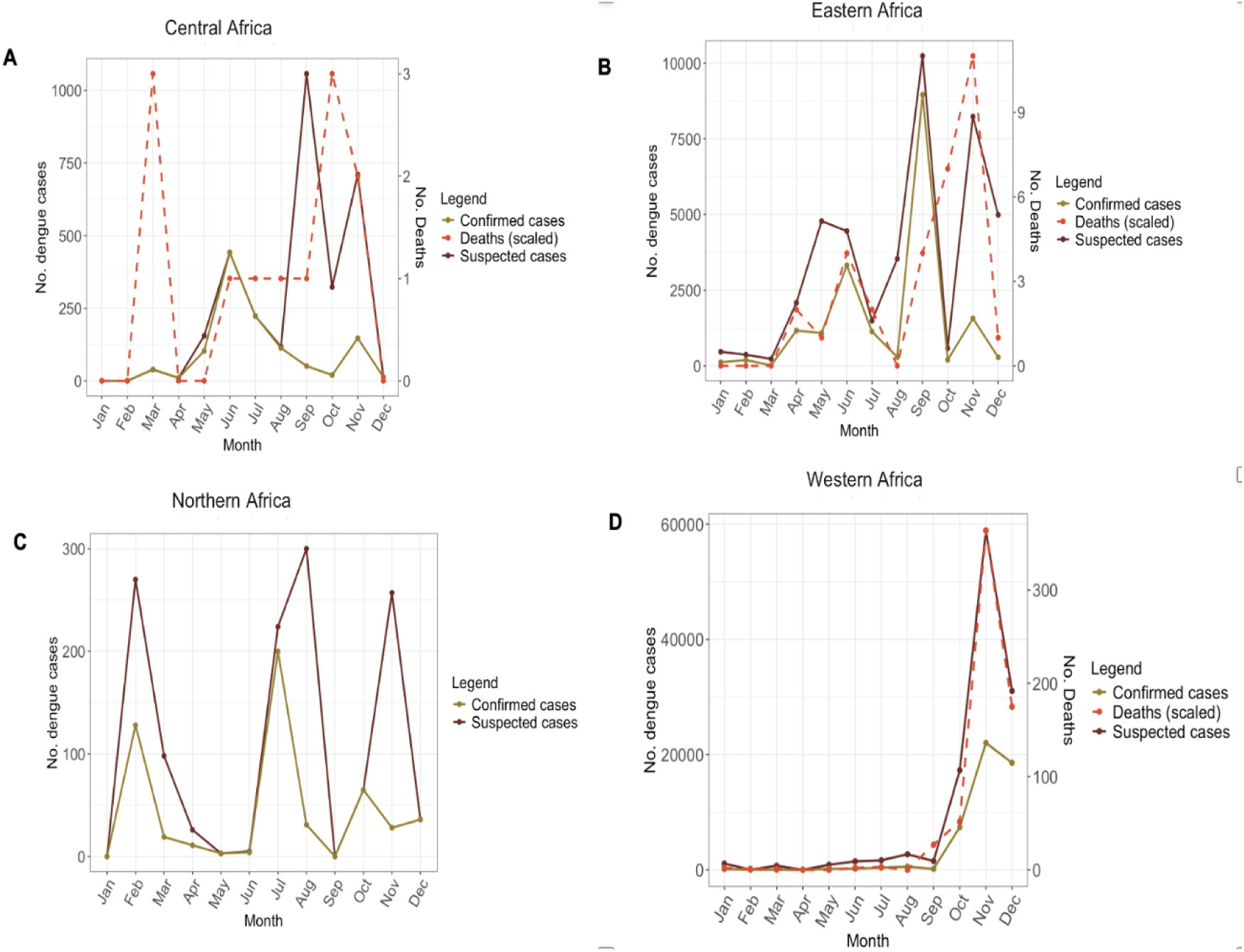
The seasonality of dengue transmission across regions based on data available from 2017–2023. A. Central Africa, B. Eastern Africa, C. Northern Africa and D. Western Africa.

Central and Western Africa experience persistent suitability (index P > 1) for dengue transmission between April and November. Eastern Africa exhibits two phases of transmission suitability (**Figure 6B**), that coincide with short and long rainy seasons from October to December and March to May, respectively, whereas Northern Africa exhibits transmission suitability between August to November (**Figure 6C**).

**Figure 6.**
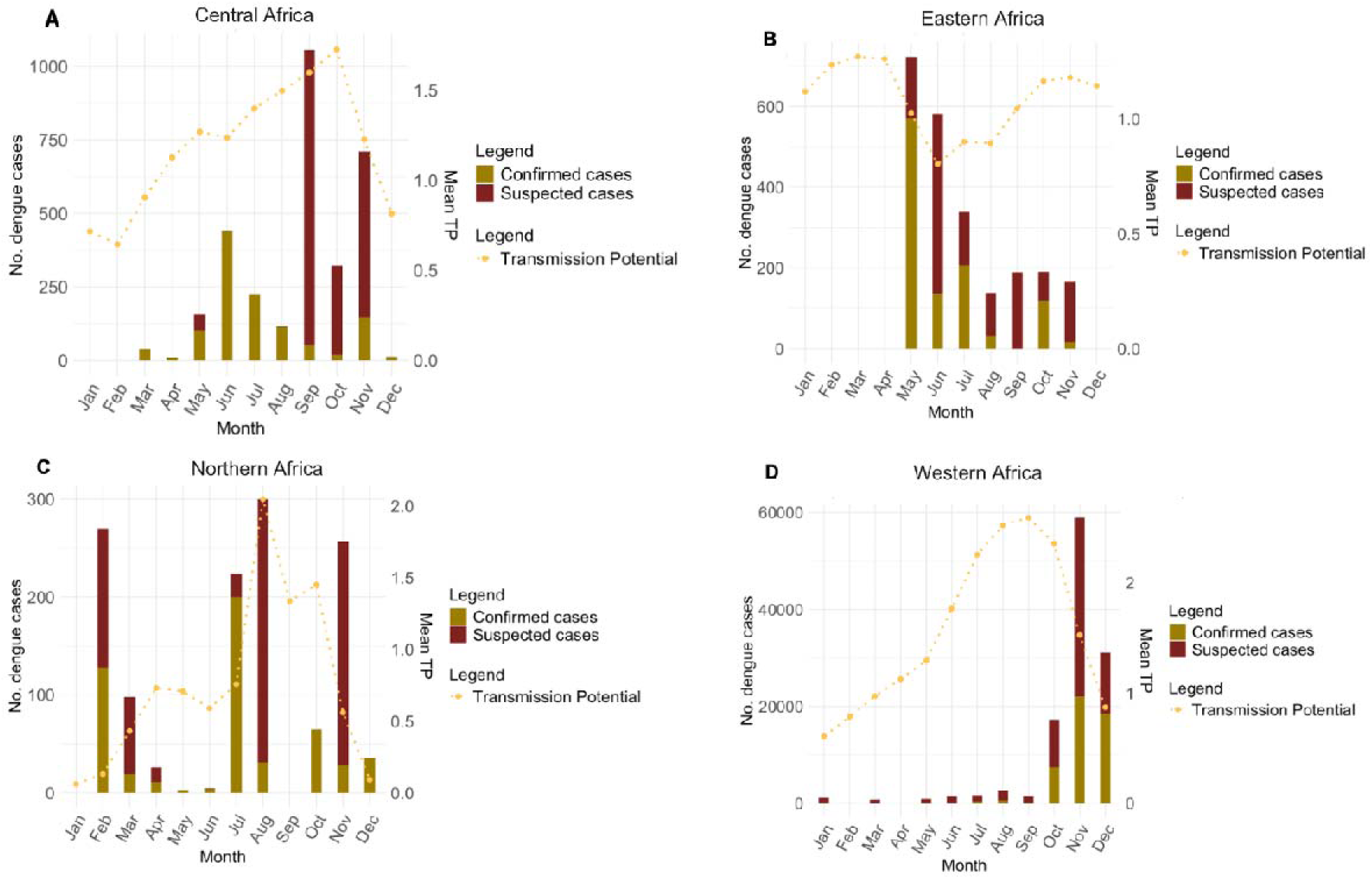
Dengue transmission potential (TP) across regions based on data available from 2017–2023. A. Central Africa B. Eastern Africa C. Northern Africa and D. Western Africa.

### 3.7 Increasing trend in the number of predicted dengue cases across Africa

The negative binomial model predicted a rising trend in the number of dengue cases in all African regions for each passing year (**Figure 7**) with the growth rate exceeding 30% in West Africa (**Table 2**). There were limited cases from Southern Africa that could be included in the model.

**Figure 7.**
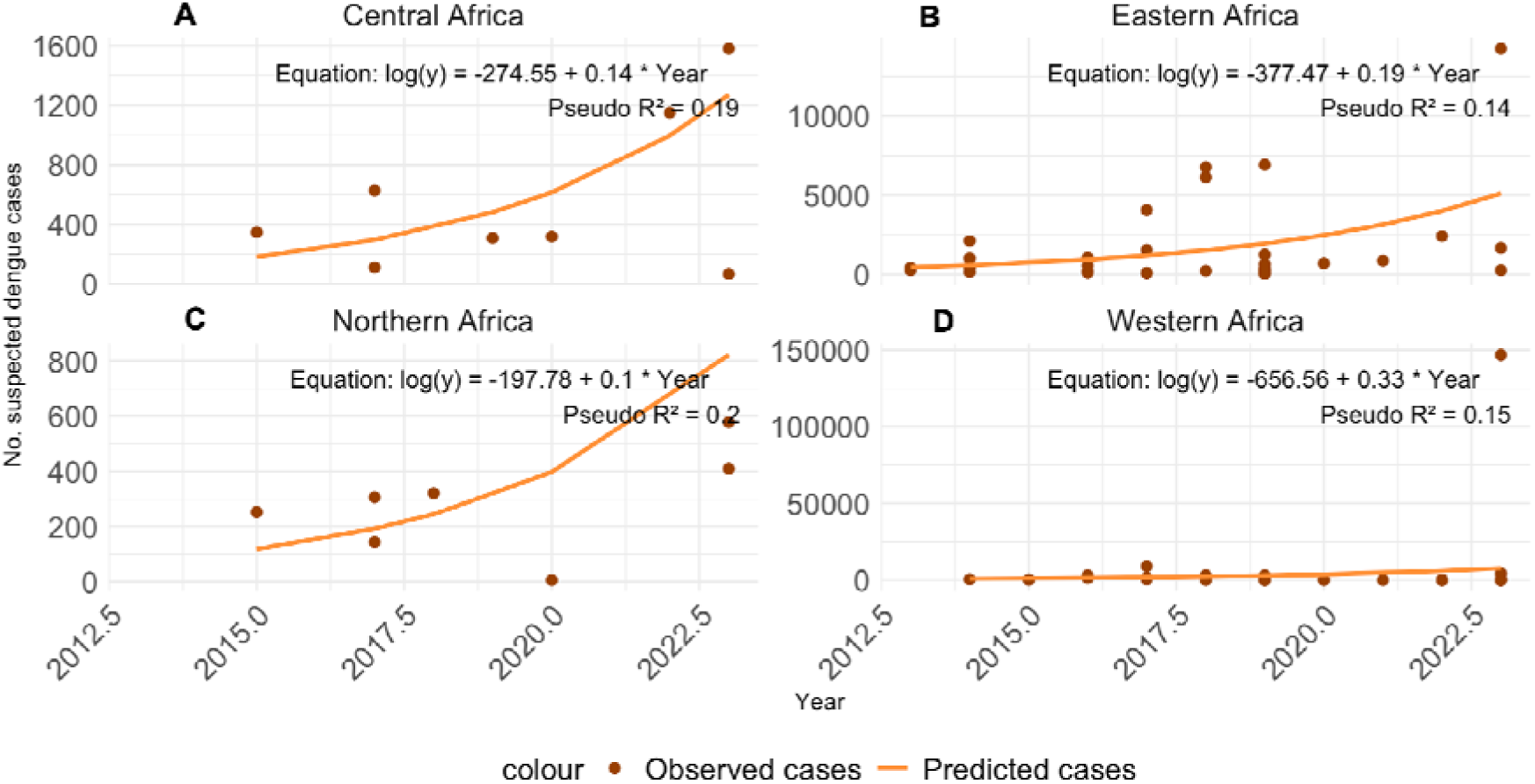
Predicted dengue cases across regions based on a negative binomial model using data reported from 2013–2023. A. Central Africa B. Eastern Africa, C. Northern Africa and D. Western Africa.

**Table 2.** Growth of predicted dengue cases in Africa regions according to negative binomial model using data available from 2013–2023.

| Region | Year coefficient ( ) | Multiplicative factor ( ) | Growth rate (%) /year |
| --- | --- | --- | --- |
| Western Africa | 0.33 | 1.39 | 39 |
| Eastern Africa | 0.19 | 1.21 | 21 |
| Central Africa | 0.14 | 1.15 | 15 |
| Northern Africa | 0.10 | 1.11 | 11 |

## 4. Discussion

Dengue represents a significant public health threat in Africa. However, the non-specific clinical presentation of the disease, which resembles malaria and other febrile illnesses such as yellow fever and chikungunya, limits better detection, reporting and understanding of the disease burden. Additionally, there are limited health resources for surveillance and timely detection [82]. This review presents the first systematic analysis to define the epidemiological trends of dengue disease burden in Africa and associated territories from 2013 to 2023. The findings of this review can inform the strengthening of intervention strategies to reduce morbidity and mortality of dengue in Africa.

The spatial analysis reveals disparities in the quantity and distribution of suspected dengue cases across Africa (**Figure 2**). This observation may indicate gaps in epidemiological surveillance and case reporting, given that tropical regions in Africa shares similar vector ecologies and transmission indices. Overall, West Africa was responsible for more than two-thirds of confirmed dengue cases and one-third of surveillance reports, indicating increasing transmission activities and improved case reporting in this region.

Burkina Faso recorded the highest burden of cases per 100,000 population (**Figure 3)**. In 2023, the country accounted for more than 80% of confirmed cases and deaths. These estimates are consistent with the World Health Organization’s surveillance report on health emergency situations [83]. The impact of climate hazards on the distribution of vectors and extensive international travels of infected individuals from endemic countries are likely to exacerbate dengue transmission through spillover events and introductions, respectively [84,85].

There are several probable factors that may have contributed to dengue transmission in Western Africa over the past decade. Several studies have demonstrated that the abundance of *Aedes aegypti* breeding habitats, particularly waste tyres, was a significant factor for transmission, particularly in peri urban centres [86,87]. Other studies have revealed that dengue outbreaks in Western Africa are driven by the presence of abundant infected *Aedes* vectors [88]. In 2023, Ouédraogo and others reported the presence of a high number of immature *Aedes aegypti* vectors in the handwashing stations that were constructed in public areas during the COVID-19 pandemic in Burkina Faso [89].

Despite a significant increase in the number of dengue cases, case fatality rate remains below 1% in Africa (**Figure 3**), compared to 3%–10% in Asia [90]. Given the continuous circulation of multiple DENV serotypes within the same region (**Figure 4**), more cases of severe dengue were expected due to lack of cross immunity.

However, it is possible that limited diagnostic capabilities, and under-reporting due to misdiagnosis with other febrile illnesses, such as malaria are contributing to low prevalence [91]. Further, genetic evidence from global ancestral analysis suggests that African descendants may be protective against dengue haemorrhagic phenotype [92].

The seasonality of dengue transmission in Africa shows regional differences (**Figure 5**), with Central and Eastern Africa experiencing long transmission seasons that coinciding with rainy seasons from May to November in Central Africa (**Figure 5A**) and from April to May and November to December in Eastern Africa (**Figure 5B**). The erratic transmission pattern observed in Northern Africa (**Figure 5C**), may be attributed to various factors including the storage of water in open containers [93,94] and heavy rainfall [95], that attract *Aedes* mosquitoes. Moreover, there are inter-regional migrations of people from endemic countries and extensive intra-regional trade activities that play a significant role in the transmission of dengue [93]. Western Africa exhibits a distinct high transmission season between October and December. These observations are consistent with the results from previous studies conducted in this region [96,97].

From April to November, Central and Western Africa (**Figure 6A and 6D**), exhibit long transmission suitability periods (Index P > 1.0) that correlate with the annual rainfall seasons [98,99]. In contrast, high transmission potential in Eastern Africa (**Figure 6B**), correlates with two rainy seasons from September to December and from March to May [100]. The low number of dengue cases reported in Northern Africa could be due to a short transmission suitability period between August to October. In general, West Africa experiences the highest transmission suitability (Index P > 2.0) between July and November in comparison to other regions. The prolonged period of transmission suitability may have contributed to the rise of Western Africa as a hotspot of dengue transmission.

The negative binomial model predicted an increasing trend of suspected dengue cases across Central, Eastern and Western Africa (**Figure 7**), with a growth rate exceeding 30% in West Africa (**Table 2**). Results from previously described climate suitability models indicate that these regions will experience significant growth in dengue incidences over the coming decades [101]. These findings will aid to inform healthcare policy and practices in Africa to enhance surveillance and implement effective interventions to prevent ongoing dengue transmission.

## Limitations

The results of this review are subject to several limitations. First, confirmed case counts may be underestimated due to the application of different case definitions. Second, limited dengue surveillance and case reporting between 2020–2022 period due to COVID-19 pandemic introduces bias in estimating dengue burden. Third, confirmed cases among travellers returning from African countries and territories were not included. Therefore, dengue burden estimates reported in this report should be interpreted with caution.

## 5. Conclusions

Over the past decade, there has been a rise in the number of confirmed dengue cases, particularly in West Africa. The persistent presence of multiple DENV serotypes within the same region increases the likelihood of severe dengue due to the lack of cross-immunity. It is important to strengthen surveillance and implement region-specific interventions to prevent future dengue outbreaks. We advocate further research for understanding the evolution and transmission dynamics of the specific dengue virus lineages in Africa.

## Institutional review board statement

No ethical clearance because this is a review of published research.

## Informed consent statement

Not applicable

## Data availability statement

All the relevant data are contained within the manuscript. Any additional data is made available through Mendeley data repository accessible at https://data.mendeley.com/my-data/.

## Authors’ contributions

**Gaspary O. Mwanyika**: Conceptualization, data curation, formal analysis, methodology, validation, writing-original draft, writing-reviewing & editing. **Abdualmoniem O. Musa**: Data curation and writing-reviewing & editing. **Jenicca Poongavanan**: Methodology, writing-reviewing & editing. **Monika Moir**: Methodology, Writing-reviewing & editing. **Gr**a**eme Dor**: Writing-review & editing. **Eduan Wilkinson**: Writing-reviewing & editing. **Cheryl Baxter:** Methodology, project administration, writing-original draft, writing-reviewing & editing. **Tulio de Oliveira:** Resources, funding acquisition, project administration, validation, writing-reviewing & editing. **Houriiyah Tegally**: Methodology, data curation, validation, writing-original draft and writing-reviewing & editing.

## Acknowledgements

We acknowledge the World Health Organization for Africa (#WHOAfro) and Africa Centers for Disease Control and Prevention (#AfricaCDC) that made epidemiological surveillance reports for different countries accessible. We also express our gratitude to all the members of the Climate Amplified Disease Epidemics (#CLIMADE) consortium for their valuable contributions in this review.

## Supplementary materials

Mwanyika, Gaspary (2024), “S1.Advanced search strategy for PubMed/Medline databases”, Mendeley Data, V1, https://data.mendeley.com/datasets/zf7x7wy7xg/1

Mwanyika, Gaspary (2024), “S2. Excluded articles for specific reasons.”, Mendeley Data, V1, https://data.mendeley.com/datasets/3gjypt9yvs/1

Mwanyika, Gaspary (2024), “S3. PRISMA 2020 checklist”, Mendeley Data, V1, https://data.mendeley.com/datasets/hx4r4jywmb/1

Mwanyika, Gaspary (2024), “S4. R programming codes”, Mendeley Data, V1, https://data.mendeley.com/datasets/d9kyhgsx2m/1

Mwanyika, Gaspary (2024), “S5. Transmission potential (Index P) data”, Mendeley Data, V1, https://data.mendeley.com/datasets/cv9c4gzdy9/1

